# Pretreatment bone turnover status and long-term bone mineral density response to denosumab

**DOI:** 10.64898/2026.10.20.26366058

**Authors:** Tomoyuki Asada, William Richardson, Soji Tani, Mahoko Ishikawa, Philip Varnadore, Tomoko Towatari, Peter Passias, Yoshifumi Kudo, Benjamin Alman, Koji Ishikawa

## Abstract

**Summary:** Pretreatment low-turnover status identified patients with attenuated total hip BMD gain after denosumab. Integrated assessment of bone resorption and formation markers may help stratify long-term hip response.

**Introduction:** Denosumab increases bone mineral density (BMD) and reduces fracture risk, but BMD response varies among patients. Given its potent antiresorptive action, denosumab may produce greater BMD gains in patients with higher pretreatment bone turnover; however, its relationship with long-term BMD response remains unclear. We developed a turnover index (TI) integrating bone resorption and formation markers and evaluated its association with BMD response trajectories up to 5 years after denosumab initiation.

**Methods:** This retrospective observational study included 230 patients with primary osteoporosis who initiated denosumab between January 2014 and January 2025. TI was calculated as the mean of standardized log-transformed TRACP-5b and total P1NP values. Patients were classified into high-, intermediate-, and low-turnover groups according to TI quartiles. Lumbar spine and total hip BMD changes were analyzed using mixed-effects models.

**Results:** The cohort included 52 high-turnover, 114 intermediate-turnover, and 64 low-turnover patients. In both unadjusted and multivariable-adjusted models, BMD gain trajectories differed significantly among TI groups at the lumbar spine and total hip (all p < 0.001). Adjusted BMD gains showed a stepwise gradient across TI groups. Five-year adjusted lumbar spine BMD gains were 20.31%, 17.01%, and 12.16% in the high-, intermediate-, and low-turnover groups, respectively. Corresponding total hip BMD gains were 10.56%, 6.76%, and 2.49%, with significantly lower gains in the low-turnover group compared to the other two groups.

**Conclusions:** Baseline bone turnover status was associated with long-term BMD response to denosumab. Low pretreatment turnover may identify patients at risk for attenuated BMD gain during denosumab treatment.

## Introduction

Denosumab is a fully human monoclonal antibody that inhibits receptor activator of nuclear factor-κB ligand (RANKL). It acts as an antiresorptive agent by rapidly suppressing bone turnover markers (BTMs) after treatment initiation, increasing bone mineral density (BMD) at both the lumbar spine and total hip, and reducing vertebral and hip fracture risk in patients with osteoporosis. [1, 2] Long-term studies have shown that BMD gains can continue for up to 10 years, with a favorable long-term safety profile. [3]

BMD response to denosumab is clinically relevant because treatment-related gains are associated with reductions in fracture risk. [4] Total hip BMD may be of particular interest because lower hip BMD and accelerated hip bone loss have also been associated with increased mortality. [5, 6] Therefore, identifying patients who are at risk for suboptimal BMD response before treatment initiation may help inform osteoporosis management.

Baseline bone turnover status may be one factor associated with variability in denosumab response. Clinical guidelines and expert recommendations have recognized BTMs as useful indicators of skeletal remodeling activity and early treatment response, [7] and bone turnover assessment has also been proposed as a potential tool for guiding osteoporosis management. [8, 9] However, the role of baseline bone turnover status in anticipating treatment response remains uncertain, despite the physiological rationale that patients with higher baseline turnover may respond more strongly to antiresorptive therapy. Clinical evidence linking pretreatment bone turnover status to long-term BMD response to denosumab remains limited. Although previous studies have linked baseline or early changes in individual BTMs with BMD response during denosumab treatment, [10–12] these studies have focused on single markers or relatively short-term outcomes. Therefore, evaluating whether an integrated assessment of baseline bone resorption and formation markers can stratify long-term BMD response to denosumab is of clinical interest.

Previous studies have integrated bone formation and resorption markers to characterize skeletal remodeling as a single metric in other clinical settings. The ratio of TRACP-5b to P1NP has been associated with nonunion following spinal fusion surgery. [13] Similarly, the anabolic window and related uncoupling indices have been used to characterize the balance between bone formation and resorption during anabolic therapy. [14–16] Drawing on this framework, we developed a turnover index (TI) integrating the bone resorption marker TRACP-5b and the bone formation marker P1NP. We investigated whether baseline TI was associated with 5-year BMD gains after denosumab initiation in patients with primary osteoporosis.

## Materials and Methods

### Study Design and Patient Selection

This retrospective observational study included patients with primary osteoporosis treated with denosumab. The study was approved by the institutional ethics committee and was conducted in accordance with the principles of the Declaration of Helsinki. The requirement for written informed consent was waived because of the retrospective nature of the study. Patients who initiated denosumab treatment between January 2014 and January 2025 were screened. During the initial screening process, patients with secondary osteoporosis, poorly controlled thyroid disease, active malignancy, or medical comorbidities that could substantially affect ambulatory status or bone quality were excluded. After this screening process, 282 patients with primary osteoporosis were considered eligible for further assessment. Among these screened patients, the following exclusion criteria were applied: (1) missing baseline DXA assessment of the lumbar spine and/or total hip, and (2) no available follow-up DXA data within 5 years after denosumab initiation. DXA follow-up data obtained within 5 years after denosumab initiation were included in the longitudinal analyses. The final analytic cohort consisted of 230 patients. (Fig. 1)

**Fig. 1.**
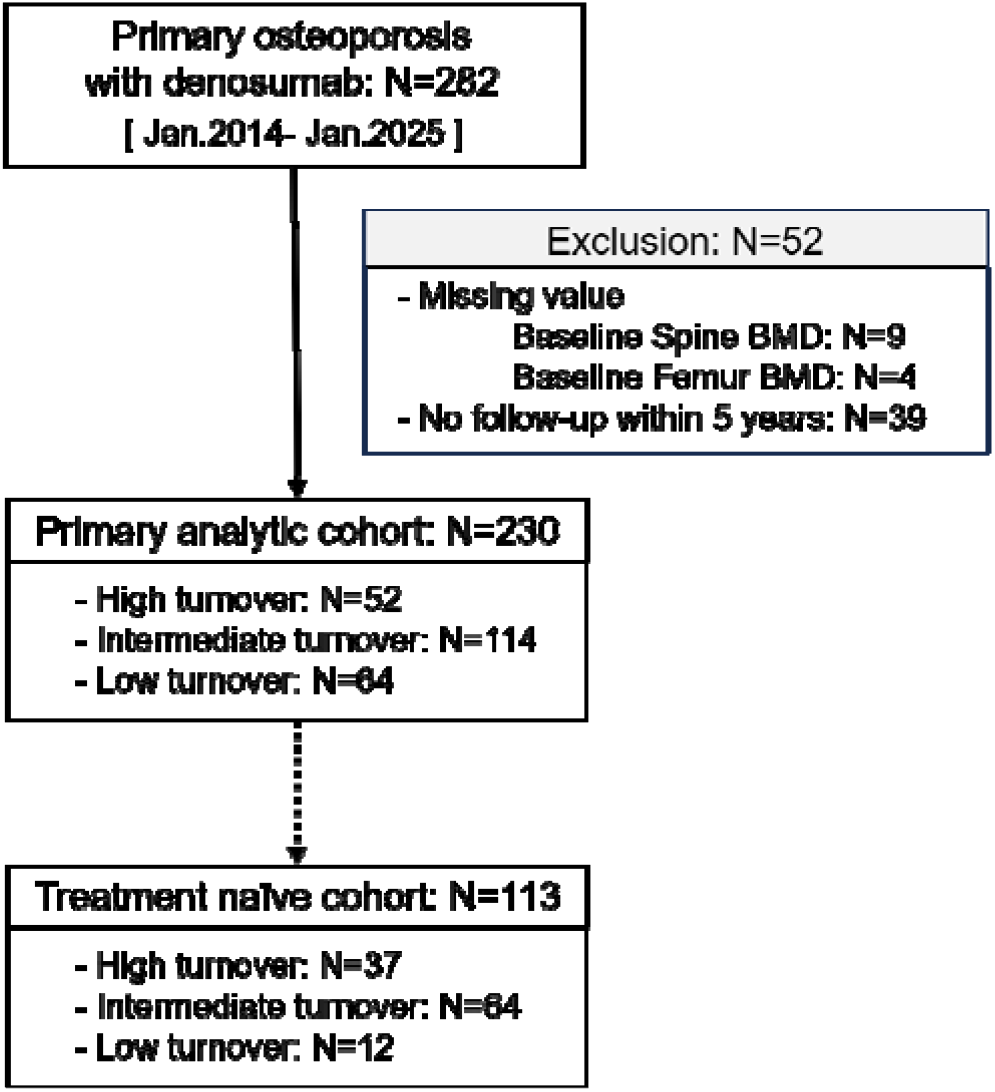
Patient flow diagram. The final analytic cohort included 230 patients with primary osteoporosis treated with denosumab.

### Data collection

Baseline demographic and clinical variables, including age, sex, BMI (kg/m^2^), current smoking status, daily alcohol consumption, and a history of fragility fracture, were collected from medical records and a questionnaire. Prior osteoporosis treatments were categorized into bisphosphonate, parathyroid hormone analogs, romosozumab, selective estrogen receptor modulators, and treatment-naïve, defined as no prior osteoporosis treatment. To address the potential influence of prior anti-osteoporosis treatment on subsequent BMD response to denosumab, analyses were conducted in the overall cohort as the primary analysis and repeated in the treatment-naïve subgroup as a sensitivity analysis.

Laboratory variables assessed before treatment included serum albumin (g/dL), calcium (mg/dL), phosphorus (mg/dL), estimated glomerular filtration rate (eGFR, mL/min/1.73 m^2^) and alkaline phosphatase (ALP, U/L). Mortality, causes of death, incident fractures during follow-up, and major osteoporotic fractures were collected from medical records.

Dual-energy X-ray absorptiometry (DXA) of the lumbar spine and total hip was performed at baseline and annually for up to 5 years after denosumab initiation. Areal BMD of the lumbar spine (L1–L4) and total hip was measured using DXA (Hologic QDR series; Hologic, Waltham, MA, USA). All DXA measurements were analyzed centrally by a radiologist. The primary longitudinal outcome was percent BMD gain from baseline (%BMD gain), calculated as 100 × (follow-up BMD − baseline BMD) / baseline BMD. Change in T-score was calculated as follow-up T-score − baseline T-score.

### Bone Turnover Markers and Turnover Index

Serum BTMs included tartrate-resistant acid phosphatase 5b (TRACP-5b; reference range in women, 120–420 mU/dL; measured using the Osteolinks® TRACP-5b® test kit, DS Pharma Biomedical Co., Ltd., Osaka, Japan) and total N-terminal propeptide of type I procollagen (total P1NP; reference range in postmenopausal women, 26.4–98.2 μg/L; measured using a total P1NP assay on an Elecsys automated analyzer, Roche Diagnostics, Switzerland). BTMs were assessed at baseline and during follow-up for up to 5 years after denosumab initiation.

Turnover index (TI) was derived as a measure of the integrated turnover activity between bone formation marker (total P1NP) and bone resorption marker (TRACP-5b). Because BTMs showed skewed distributions, TRACP-5b and P1NP were first log-transformed and then standardized to z-scores (zTRACP-5b and zP1NP, respectively). The TI was defined as the mean of these two standardized values.

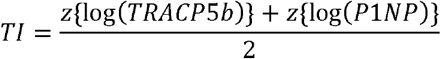

Higher TI values represented higher bone turnover. For categorical analyses based on TI, patients were classified into three TI groups according to baseline TI quartiles. Patients in the highest quartile were classified as the high-turnover group (H group), those in the lowest quartile as the low-turnover group (L group), and those in the second and third quartiles as the intermediate-turnover group (I group).

### Statistical analysis

All statistical analyses were performed using R software (version 4.4.1; R Core Team, Vienna, Austria). Continuous variables were summarized as means with standard deviations or medians with interquartile ranges, as appropriate. Categorical variables were summarized as counts and percentages. Univariate comparisons across TI groups were performed using Welch’s one-way ANOVA for continuous variables and chi-square test for categorical variables. For BTMs, Kruskal–Wallis test was used for comparison given the non-normal distribution.

Multivariable linear mixed-effects models were used to investigate the association between baseline TI groups and %BMD gain and absolute change in T-score over 5 years after denosumab initiation. The models included all available repeated measurements without imputation and assumed that missing follow-up outcome data were missing at random conditional on observed outcomes and covariates included in the models. Given a possible non-linear relationship between treatment duration and %BMD gain, follow-up time was modeled using natural cubic splines with 3 degrees of freedom, and an interaction term between baseline TI group and follow-up time was included to estimate the associations. Models were constructed separately for lumbar spine BMD and total hip BMD. All models were adjusted for age, sex, BMI, eGFR, baseline BMD or T-score at each site, prior osteoporosis treatment, and type of vitamin D supplementation during denosumab treatment in the overall cohort. The overall interaction was evaluated using Type III analysis of variance with Satterthwaite’s approximation for degrees of freedom. Models included patient-specific random intercepts and random slopes for follow-up time. Sensitivity analysis was conducted with the treatment-naïve cohort to minimize the influence from prior treatment. Multivariable models with the treatment-naïve cohort were also adjusted for the same covariates without prior treatment.

Adjusted changes from baseline at 1, 2, and 5 years were estimated as contrasts between each follow-up time point and baseline within each TI group. Between-group differences were evaluated using contrasts of these within-group changes. P values were adjusted using the Holm method for the three pairwise comparisons within each time point.

Longitudinal changes for BTMs were also analyzed using linear mixed-effects models of natural-log-transformed ratios of follow-up to baseline values. Models included follow-up time, TI group, and their interaction; were adjusted for age, sex, BMI, eGFR, prior osteoporosis treatment, and type of vitamin D supplementation; and included a patient-specific random intercept. Estimates were back-transformed and expressed as percentage changes from baseline. A two-sided p-value <0.05 was considered statistically significant.

## Results

### Baseline patient characteristics

Of the 230 patients, 52 were classified into the high-turnover group, 114 into the intermediate-turnover group, and 64 into the low-turnover group, and 113 were treatment-naïve. BMD measurements were available for 213, 189, and 113 patients at 1, 2, and 5 years, respectively. Among the treatment-naïve patients, BMD measurements were available for 103, 87, and 60 patients at 1, 2, and 5 years, respectively.

Age and sex distributions and baseline lumbar spine BMD and T-score were similar across the TI groups.

Differences were observed in BMI, fracture history, prior treatment, and baseline total hip BMD and T-score, as well as in several biochemical parameters (Table 1). The overall pattern of baseline characteristics across the TI groups was similar in the treatment-naïve subgroup and the overall cohort (Supplementary Table S1). Figure 2 illustrates the baseline distribution of TRACP-5b and total P1NP according to the quartile-based TI group classification. The cutoff values defining the high- and low-turnover groups are provided in Supplementary Table S2.

**Fig. 2.**
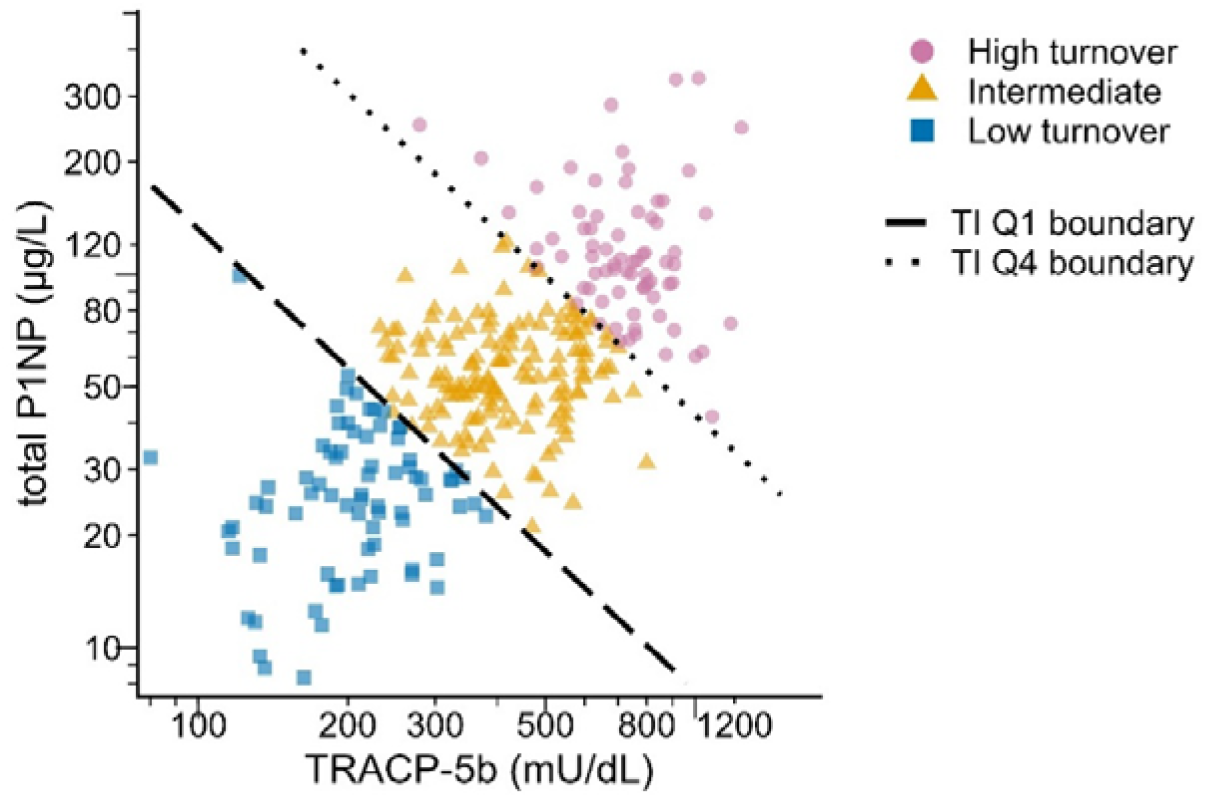
Baseline TRACP-5b and total P1NP distributions by TI group. Scatter plot showing the baseline distribution of TRACP-5b and total P1NP according to TI group. TI was calculated as the mean of standardized log-transformed TRACP-5b and total P1NP values. Patients in the highest quartile were classified as the high-turnover group, those in the lowest quartile as the low-turnover group, and those in the second and third quartiles as the intermediate-turnover group. Dashed and dotted lines indicate TI quartile boundaries.

### Longitudinal changes in BTMs over 5 years by TI group

Both TRACP-5b and P1NP decreased rapidly after denosumab initiation, with the greatest reductions observed in the high-turnover group and the smallest reductions in the low-turnover group (Fig. 3a-b). In multivariable mixed-effects models, the longitudinal suppression pattern differed significantly according to TI group for both P1NP and TRACP-5b (both p < 0.001 for the interaction between follow-up time and TI group).

**Fig. 3.**
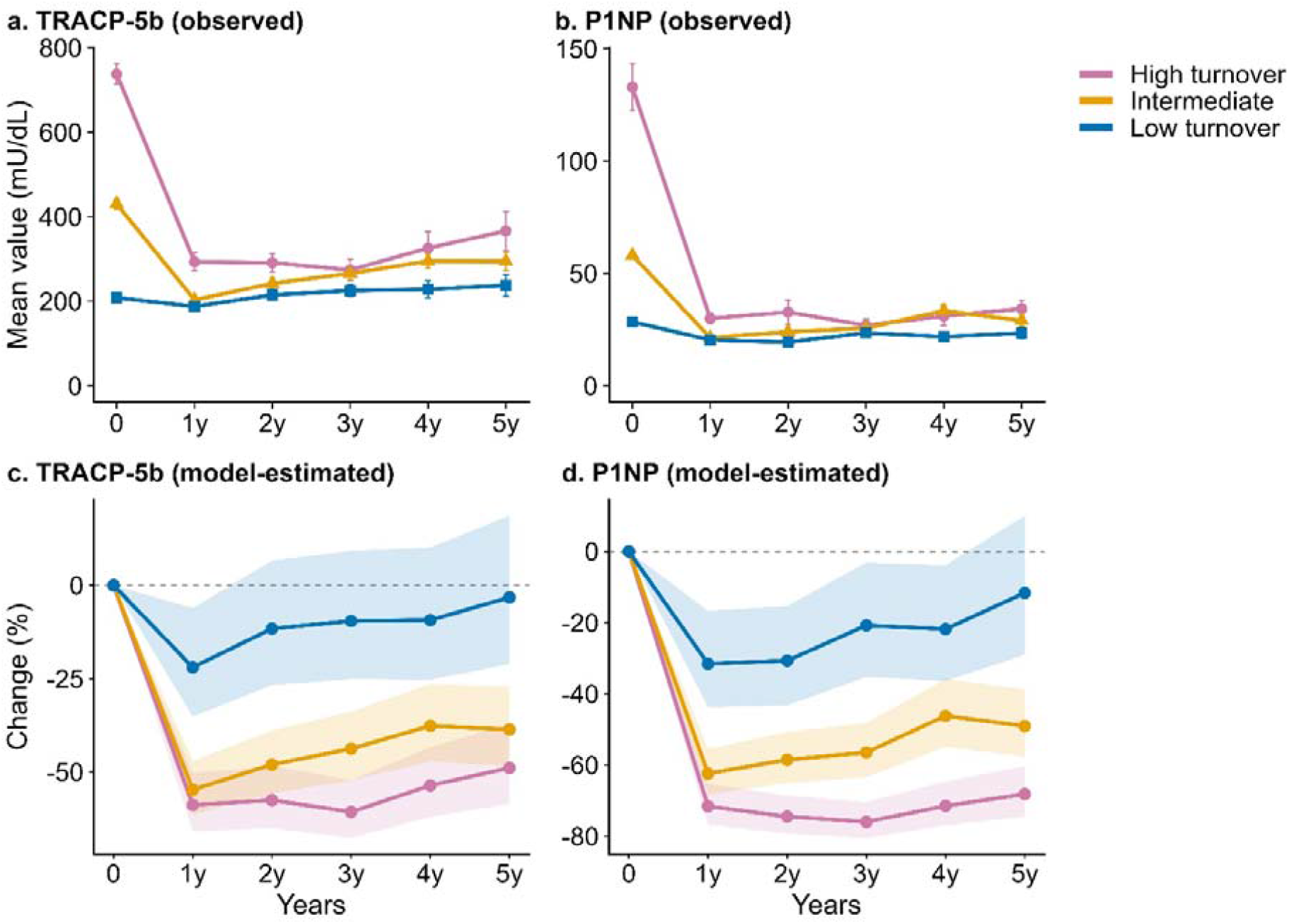
Longitudinal changes in BTMs according to baseline TI group. Observed mean values of TRACP-5b (a) and total P1NP (b) are shown from baseline to 5 years after denosumab initiation according to baseline TI group. Error bars indicate standard errors. Figure c and d show adjusted model-estimated percentage changes derived from natural-log-transformed within-patient follow-up-to-baseline ratios; shaded areas indicate 95% confidence intervals.

In model-estimated percent changes, TRACP-5b remained reduced at 5 years by 48.9%, 38.6%, and 3.2% in the high-turnover, intermediate, and low-turnover groups, respectively. Corresponding reductions in P1NP at 5 years were 68.2%, 49.1%, and 11.6%, respectively (Fig. 3c-d). These findings suggest that patients with higher TI showed more pronounced and sustained BTM suppression after denosumab, whereas those in the low-turnover group showed relatively limited additional suppression.

### Association between TI group and lumbar spine BMD response

In the multivariable mixed-effects models adjusted for age, sex, BMI, eGFR, baseline DXA, prior treatment, and type of vitamin D supplementation, TI group was significantly associated with the trajectories of both %BMD gain and change in T-score (Year×Group interaction term in both of the outcomes, p < 0.001; Supplementary Table S3). Adjusted trajectories demonstrated a graded pattern, with the greatest improvement in the high-turnover group, intermediate improvement in the intermediate group, and the smallest improvement in the low-turnover group (Fig. 4). Adjusted estimates and pairwise comparisons at each time point are presented in Table 2. At 5 years, adjusted spine %BMD gains were 20.31±1.38%, 17.01±0.95%, and 12.16±1.21% in the high-turnover, intermediate, and low-turnover groups, respectively. At 5 years, percent BMD gain was greater in the high-turnover group than in the intermediate- and low-turnover groups (p = 0.047 and p < 0.001, respectively) and was greater in the intermediate group than in the low-turnover group (p = 0.003). Corresponding changes in T-score were 1.293±0.106, 1.110±0.073, and 0.892±0.094, respectively. For T-score change, the high-turnover group showed a significantly greater increase than the low-turnover group at 5 years (p = 0.015).

**Fig. 4.**
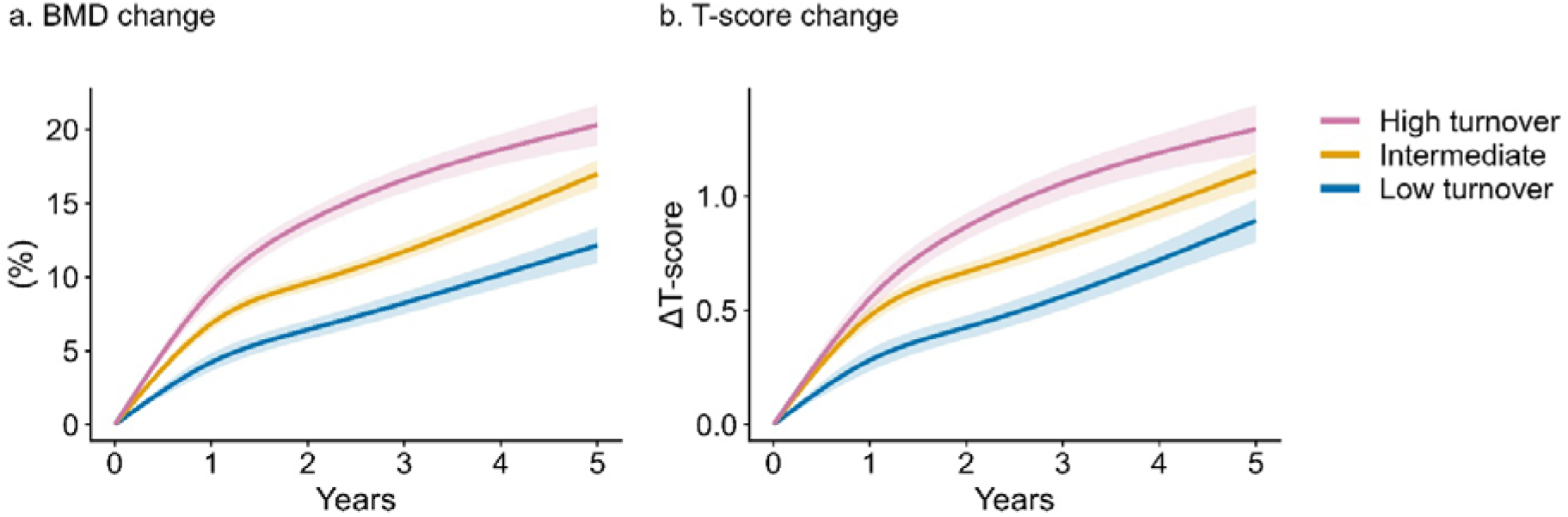
Adjusted trajectories of lumbar spine DXA outcomes according to baseline TI group. Adjusted trajectories of percent BMD gain from baseline (a) and change in T-score (b) were estimated using multivariable linear mixed-effects models with natural cubic splines for follow-up time. Models were adjusted for age, sex, BMI, eGFR, baseline lumbar spine DXA value or T-score, prior osteoporosis treatment status, and type of vitamin D supplementation during denosumab treatment. Shaded areas indicate standard errors.

The sensitivity analysis in the treatment-naïve cohort included 113 patients, consisting of 37 patients in the high-turnover group, 64 in the intermediate group, and 12 in the low-turnover group (Supplementary Table S1). Longitudinal lumbar spine DXA responses in this cohort showed a consistent pattern with the primary analysis. TI group showed significantly different BMD trajectory for both %BMD (p < 0.001) and change in T-score (p = 0.002). Adjusted trajectories also showed graded separation of BMD gain according to TI group (Supplementary Figure S1). At 2 years, estimated percent BMD gain increased in a graded manner across the high-turnover, intermediate-, and low-turnover groups (14.25%, 10.37%, and 7.11%, respectively). Gains were greater in the high-turnover group than in both the intermediate- and low-turnover groups

(Holm-adjusted p = 0.002 for both comparisons; Supplementary Table S4). There was no statistically significant differences between intermediate- and low-turnover groups at year 5 time point.

### Association between TI groups and total hip BMD response

In the both unadjusted and multivariable model for total hip, TI groups showed significantly different adjusted trajectories of percent BMD gain and T-score change over 5 years (overall interaction p < 0.001 and p = 0.001, respectively; Supplementary Table S5). At each evaluated time point, percent BMD gain and T-score improvement were lower in the low-turnover group than in the high- and intermediate-turnover groups, whereas no significant differences were observed between the high- and intermediate-turnover groups (Fig. 5). At 1 year after denosumab initiation, adjusted %BMD gains were 3.65%, 2.55%, and 0.94% in the high-turnover, intermediate, and low-turnover groups, respectively. This trend was also consistent at 5 years, showing 10.56%, 6.76%, and 2.49%. Corresponding changes in T-score were also consistent. (Table 3)

**Fig. 5.**
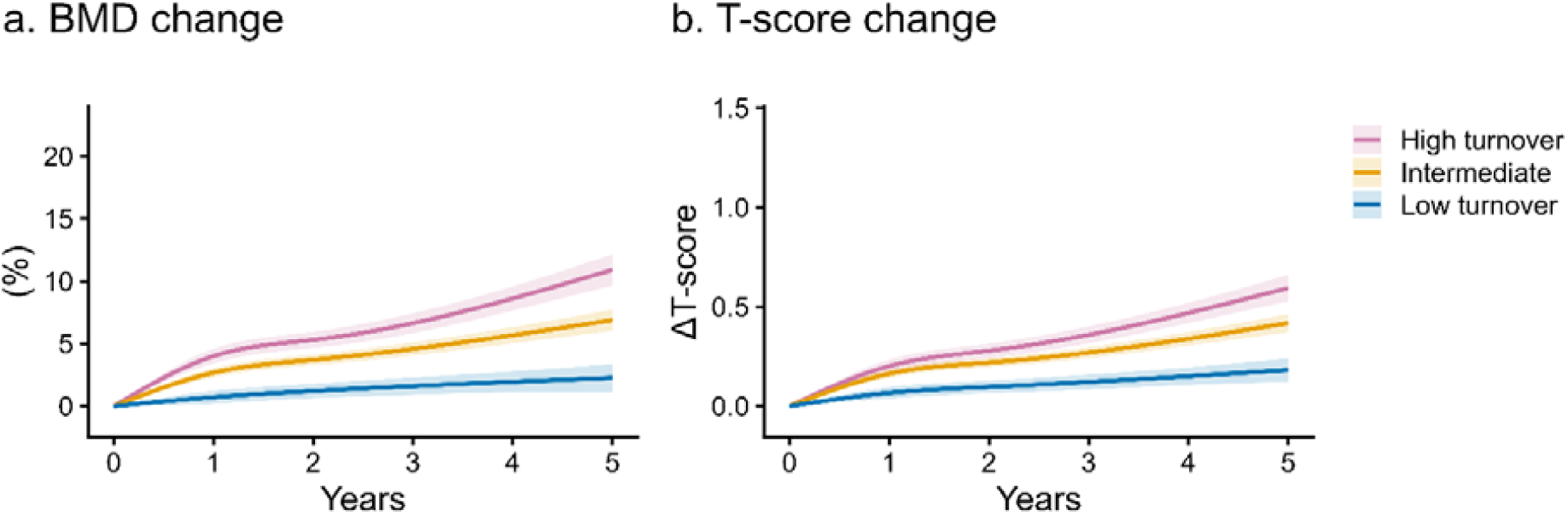
Adjusted trajectories of total hip DXA outcomes according to baseline TI group. Adjusted trajectories of percent BMD gain from baseline (a) and change in T-score (b) were estimated using multivariable linear mixed-effects models with natural cubic splines for follow-up time. Models were adjusted for age, sex, BMI, eGFR, baseline total hip DXA value or T-score, prior osteoporosis treatment status, and type of vitamin D supplementation during denosumab treatment. Shaded areas indicate standard errors.

In the treatment-naïve cohort, BMD gains tended to remain greater in the high- and intermediate-turnover groups than in the low-turnover group, although between-group differences did not reach statistical significance. (Supplementary Fig. S2, Supplementary Table S6)

### Mortality and fracture events during follow-up

Death occurred in 8 patients (15.4%) in the high-turnover group, 15 (13.2%) in the intermediate group, and 6 (9.4%) in the low-turnover group, with no statistically significant difference across groups (p = 0.61). All death were not directly related to denosumab treatment. The distribution of causes of death was heterogeneous, with pneumonia being the most frequent cause in the high-turnover groups. The number of incident fractures during follow-up was also similar across groups, and the proportion of patients with any follow-up fracture did not differ by TI group. Major osteoporotic fractures occurred in 4 patients (7.7%) in the high-turnover group, 10 (8.8%) in the intermediate group, and 7 (10.9%) in the low-turnover group, without a statistically significant difference across groups (p = 0.82). (Supplementary Table S7)

## Discussion

This study developed a TI integrating bone resorption and formation markers and evaluated whether baseline TI could stratify long-term BMD responses to denosumab in patients with osteoporosis. Patients in the low-turnover group showed attenuated BMD gains. These findings suggest that pretreatment bone turnover status may help identify patients who are likely to have a lower BMD response to denosumab.

Bone turnover assessment has been proposed as a potential tool for guiding osteoporosis management, but the clinical utility of individual baseline BTMs remains uncertain. [8] Previous studies have linked individual baseline BTMs or their early changes to BMD gains during denosumab treatment. [10–12, 17] For example, in the DATA study, higher pretreatment P1NP and CTX levels were each associated with greater 2-year BMD gains among patients receiving denosumab. [11] However, whether combined assessment of bone formation and resorption can stratify long-term treatment response remains unclear. Our findings address this gap by showing that TI-defined baseline turnover groups were associated with distinct 5-year BMD response trajectories during denosumab treatment.

Although bone resorption and formation are biologically coupled and their markers generally change in parallel, their relative levels may vary among individuals, and the two processes may become partially uncoupled under pathological conditions, during osteoporosis treatment, or when switching therapies. [18] Ratio-based approaches have also been proposed to characterize the balance between bone formation and resorption. However, these approaches may not capture overall turnover intensity, because similar ratios can occur when both markers are low or when both are high. [18, 19] By integrating the standardized levels of both markers rather than their ratio, TI was designed to capture overall pretreatment turnover intensity. Our findings suggest that an integrated assessment of formation and resorption may provide additional insight into baseline turnover status in relation to long-term BMD response to denosumab.

Patients in the low-TI group may represent a distinct low-turnover phenotype that is not solely attributable to previous osteoporosis treatment. Although prior treatment differed across TI groups, the association between low TI and attenuated BMD response persisted after adjustment for prior treatment and other relevant baseline characteristics and was also observed in the treatment-naïve subgroup. This raises the possibility that low TI reflects underlying biological differences in skeletal turnover that may influence the magnitude of BMD response to denosumab. However, the small number of treatment-naïve patients with low TI limits the strength of this interpretation and warrants confirmation in larger cohorts.

Longitudinal changes in BTMs may help explain the differences in long-term BMD response among TI groups. TRACP-5b and P1NP were markedly suppressed in the high- and intermediate-turnover groups, whereas the low-turnover group showed little additional suppression. Although the absolute differences in BTMs narrowed soon after denosumab initiation, the relative ordering of the groups persisted, indicating that longitudinal BTM profiles continued to reflect pretreatment turnover status. This persistent difference in turnover patterns may have contributed to the distinct BMD response trajectories among the TI groups.

Several limitations should be acknowledged. First, because of the retrospective observational design, residual confounding cannot be excluded. Second, TI groups were defined using cohort-specific quartiles, which may limit the generalizability of the group definitions. Third, the treatment-naïve sensitivity analysis was limited by the small number of patients. Finally, racture outcomes could not be robustly analyzed because of the limited number of events.

## Conclusion

In patients with primary osteoporosis treated with denosumab, a pretreatment TI integrating bone resorption and formation markers was associated with long-term BMD response. Patients with a low-turnover group showed attenuated BMD gain over 5 years. These findings suggest that pretreatment bone turnover status may help identify patients who are less likely to achieve adequate BMD gain with denosumab. Further prospective studies are needed to validate this group and to determine whether alternative treatment strategies may improve outcomes in low-turnover patients.

## Data Availability

Data are available from the corresponding author upon reasonable request and with appropriate institutional approval.

## Acknowledgments

We thank Ayano Oyamada, Kei Gonsho, Miho Mochizuki, and Tomoko Towatari for their cooperation in collecting clinical data.

## Reference

1. Bone HG, Chapurlat R, Brandi M-L, et al (2013) The effect of three or six years of denosumab exposure in women with postmenopausal osteoporosis: results from the FREEDOM extension. J Clin Endocrinol Metab 98:4483–4492

2. Cummings SR, San Martin J, McClung MR, et al (2009) Denosumab for prevention of fractures in postmenopausal women with osteoporosis. N Engl J Med 361:756–765

3. Bone HG, Wagman RB, Brandi ML, et al (2017) 10 years of denosumab treatment in postmenopausal women with osteoporosis: results from the phase 3 randomised FREEDOM trial and open-label extension. Lancet Diabetes Endocrinol 5:513–523

4. Black DM, Bauer DC, Vittinghoff E, et al (2020) Treatment-related changes in bone mineral density as a surrogate biomarker for fracture risk reduction: meta-regression analyses of individual patient data from multiple randomised controlled trials. Lancet Diabetes Endocrinol 8:672–682

5. Cawthon PM, Patel S, Ewing SK, et al (2017) Bone loss at the hip and subsequent mortality in older men: The Osteoporotic Fractures in men (MrOS) study: Bone loss at the hip and subsequent mortality in older men. JBMR Plus 1:31–35

6. Haentjens P, Magaziner J, Colón-Emeric CS, et al (2010) Meta-analysis: excess mortality after hip fracture among older women and men. Ann Intern Med 152:380–390

7. Camacho PM, Petak SM, Binkley N, et al (2020) American Association of Clinical Endocrinologists/American College of Endocrinology clinical practice guidelines for the diagnosis and treatment of postmenopausal osteoporosis-2020 update. Endocr Pract 26:1–46

8. Nishizawa Y, Ohta H, Miura M, et al (2013) Guidelines for the use of bone metabolic markers in the diagnosis and treatment of osteoporosis (2012 edition). J Bone Miner Metab 31:1–15

9. Nishizawa Y, Miura M, Ichimura S, et al (2019) Executive summary of the Japan Osteoporosis Society Guide for the Use of Bone Turnover Markers in the diagnosis and treatment of osteoporosis (2018 Edition). Clin Chim Acta 498:101–107

10. Eastell R, Christiansen C, Grauer A, et al (2011) Effects of denosumab on bone turnover markers in postmenopausal osteoporosis. J Bone Miner Res 26:530–537

11. Tsai JN, Burnett-Bowie SM, Lee H, Leder BZ (2017) Relationship between bone turnover and density with teriparatide, denosumab or both in women in the DATA study. Bone 95:20–25

12. Etani Y, Noguchi T, Yukishima T, et al (2025) Impact of baseline PINP on the BMD increase with romosozumab, teriparatide, and denosumab in treatment-naïve primary osteoporosis: A retrospective cohort study. Bone 201:117627

13. Inose H, Yamada T, Mulati M, et al (2018) Bone turnover markers as a new predicting factor for nonunion after spinal fusion surgery. Spine (Phila Pa 1976) 43:E29–E34

14. Eastell R, Brown JP, Adler RA, et al (2025) Bone turnover markers predict changes in bone mineral density in men treated with abaloparatide: results from the abaloparatide for the treatment of men with osteoporosis (ATOM) study. J Bone Miner Res 40:315–322

15. Eastell R, Mitlak BH, Wang Y, et al (2019) Bone turnover markers to explain changes in lumbar spine BMD with abaloparatide and teriparatide: results from ACTIVE. Osteoporos Int 30:667–673

16. Lane NE, Sanchez S, Genant HK, et al (2000) Short-term increases in bone turnover markers predict parathyroid hormone-induced spinal bone mineral density gains in postmenopausal women with glucocorticoid-induced osteoporosis. Osteoporos Int 11:434–442

17. Hori M, Yasuda K, Takahashi H, et al (2022) Effects of bone turnover status on the efficacy and safety of denosumab among haemodialysis patients. Sci Rep 12:7781

18. Shieh A, Karlamangla AS, Gossiel F, et al (2025) Estimating net bone formation relative to resorption using reference bone turnover markers. J Clin Endocrinol Metab 110:e2544–e2552

19. Nakatoh S (2017) Bone turnover rate and bone formation/resorption balance during the early stage after switching from a bone resorption inhibitor to denosumab are predictive factors of bone mineral density change. Osteoporos Sarcopenia 3:45–52

