## Supplementary material for "Pretreatment bone turnover status and long-term bone mineral density response to denosumab": Tables

**Table 1. Patient demographics**

|  | High turnover | Intermediate | Low turnover | p-value |
| --- | --- | --- | --- | --- |
| n | 52 | 114 | 64 |  |
| Age (mean (SD)) | 77.7 (7.9) | 75.9 (8.2) | 77.3 (7.9) | 0.333 |
| Male, n(%) | 4 (7.7) | 10 (8.8) | 1 (1.6) | 0.162 |
| BMI (mean (SD)) | 19.9 (5.2) | 22.2 (3.6) | 21.0 (5.9) | 0.010* |
| Fracture history, n(%) | 43 (82.7) | 73 (64.0) | 40 (62.5) | 0.033* |
| Prior treatment, n(%) |  |  |  | <0.001* |
| Bisphosphonate | 1 (1.9) | 15 (13.2) | 18 (28.1) |  |
| None/VitD | 37 (71.2) | 64 (56.1) | 12 (18.8) |  |
| PTH | 12 (23.1) | 13 (11.4) | 8 (12.5) |  |
| Romosozumab | 1 (1.9) | 20 (17.5) | 26 (40.6) |  |
| SERM | 1 (1.9) | 2 (1.8) | 0 (0.0) |  |
| Smoking, n(%) | 3 (5.8) | 6 (5.3) | 2 (3.1) | 0.758 |
| Glucocorticoid use, n(%) | 0 (0.0) | 4 (3.5) | 4 (6.2) | 0.188 |
| eGFR, mL/min/1.73 m², (mean (SD)) | 65.8 (20.5) | 65.6 (16.7) | 62.3 (18.3) | 0.466 |
| Albumin, g/dL, (mean (SD)) | 4.2 (0.3) | 4.2 (0.3) | 4.2 (0.3) | 0.156 |
| Calcium, mg/dL, (mean (SD)) | 9.3 (0.4) | 9.3 (0.4) | 9.4 (0.5) | 0.410 |
| IP, mg/dL, (mean (SD)) | 3.7 (0.5) | 3.5 (0.4) | 3.5 (0.5) | 0.028* |
| ALP, U/L, (mean (SD)) | 332.9 (110.4) | 247.6 (113.3) | 181.4 (81.7) | <0.001* |
| TRACP-5b, mU/dL, (median [IQR]) | 739.5 [621.8, 831.0] | 407.5 [331.2, 537.0] | 207.0 [176.5, 249.8] | <0.001* |
| Total P1NP, µg/L, (median [IQR]) | 107.9 [93.9, 141.8] | 54.6 [46.8, 68.9] | 25.8 [18.8, 35.1] | <0.001* |
| Lumbar spine BMD, g/cm², (mean (SD)) | 0.735 (0.160) | 0.779 (0.147) | 0.803 (0.197) | 0.104 |
| Lumbar spine T-score (mean (SD)) | -2.265 (1.431) | -1.877 (1.312) | -1.641 (1.749) | 0.097 |
| Total hip BMD, g/cm², (mean (SD)) | 0.573 (0.116) | 0.639 (0.113) | 0.655 (0.101) | <0.001* |
| Total hip T-score (mean (SD)) | -3.012 (1.158) | -2.370 (1.138) | -2.206 (1.009) | <0.001* |

BMI, body mass index; VitD, vitamin D supplementation; PTH, parathyroid hormone analog; SERM, selective estrogen receptor modulator; eGFR, estimated glomerular filtration rate; IP, inorganic phosphorus; ALP, alkaline phosphatase; TRACP-5b, tartrate-resistant acid phosphatase 5b; P1NP, N-terminal propeptide of type I procollagen; BMD, bone mineral density; SD, standard deviation; IQR, interquartile range.

*p < 0.05

**Table 2. Adjusted lumbar spine DXA up to 5-years**

| Spine BMD |  | Adjusted mean | Difference [95% CI] | Adjusted mean | Difference [95% CI] | Adjusted mean | p-value | | |
| --- | --- | --- | --- | --- | --- | --- | --- | --- | --- |
|  | year | High turnover | H vs I | Intermediate | I vs L | Low turnover | H vs I | I vs L | H vs L |
| %BMD change | 1 | 9.04 (0.68) | 2.19 [0.59, 3.79] | 6.85 (0.45) | 2.63 [1.16, 4.09] | 4.23 (0.60) | 0.007* | <0.001* | <0.001* |
|  | 2 | 13.80 (0.73) | 4.20 [2.48, 5.92] | 9.60 (0.49) | 3.16 [1.59, 4.73] | 6.44 (0.64) | <0.001* | <0.001* | <0.001* |
|  | 5 | 20.31 (1.38) | 3.30 [0.04, 6.57] | 17.01 (0.95) | 4.85 [1.85, 7.85] | 12.16 (1.21) | 0.047* | 0.003* | <0.001* |
| ΔT-score | 1 | 0.552 (0.055) | 0.076 [-0.053, 0.205] | 0.476 (0.037) | 0.194 [0.075, 0.312] | 0.282 (0.048) | 0.250 | 0.003* | <0.001* |
|  | 2 | 0.866 (0.058) | 0.198 [0.062, 0.334] | 0.668 (0.039) | 0.242 [0.117, 0.366] | 0.426 (0.051) | 0.004* | <0.001* | <0.001* |
|  | 5 | 1.293 (0.106) | 0.183 [-0.069, 0.435] | 1.110 (0.073) | 0.218 [-0.014, 0.450] | 0.892 (0.094) | 0.154 | 0.131 | 0.015* |

Values are adjusted estimated marginal means with standard errors derived from multivariable linear mixed-effects models. Models were adjusted for age, sex, BMI, eGFR, baseline DXA value at the lumbar spine, prior osteoporosis treatment status, and type of vitamin D supplementation during denosumab treatment. Differences are presented with unadjusted 95% confidence intervals. P values were adjusted using the Holm method for the three pairwise comparisons within each time point.

BMD, bone mineral density; CI, confidence interval

*p-value < 0.05

**Table 3. Adjusted total hip DXA up to 5-years**

| Total hip BMD |  | Adjusted mean | Difference [95% CI] | Adjusted mean | Difference [95% CI] | Adjusted mean | p-value | | |
| --- | --- | --- | --- | --- | --- | --- | --- | --- | --- |
|  | year | High turnover | H vs I | Intermediate | I vs L | Low turnover | H vs I | I vs L | H vs L |
| %BMD change | 1 | 3.65 (0.68) | 1.10 [-0.01, 2.21] | 2.55 (0.57) | 1.61 [0.58, 2.64] | 0.94 (0.67) | 0.051 | 0.005* | <0.001* |
|  | 2 | 4.95 (0.75) | 1.37 [0.05, 2.69] | 3.58 (0.61) | 2.14 [0.92, 3.37] | 1.44 (0.72) | 0.042* | 0.001* | <0.001* |
|  | 5 | 10.56 (1.31) | 3.80 [0.91, 6.69] | 6.76 (0.95) | 4.27 [1.61, 6.92] | 2.49 (1.19) | 0.010* | 0.003* | <0.001* |
| ΔT-score | 1 | 0.177 (0.043) | 0.028 [-0.039, 0.096] | 0.149 (0.036) | 0.078 [0.014, 0.141] | 0.071 (0.042) | 0.411 | 0.033* | 0.030* |
|  | 2 | 0.255 (0.045) | 0.050 [-0.027, 0.127] | 0.205 (0.037) | 0.104 [0.032, 0.176] | 0.101 (0.044) | 0.201 | 0.009* | 0.003* |
|  | 5 | 0.570 (0.073) | 0.166 [0.011, 0.322] | 0.403 (0.054) | 0.217 [0.074, 0.361] | 0.186 (0.067) | 0.036* | 0.006* | <0.001* |

Values are adjusted estimated marginal means with standard errors derived from multivariable linear mixed-effects models. Models were adjusted for age, sex, BMI, eGFR, baseline DXA value at the total hip, prior osteoporosis treatment status, and type of vitamin D supplementation during denosumab treatment. Differences are presented with unadjusted 95% confidence intervals. P values were adjusted using the Holm method for the three pairwise comparisons within each time point.

BMD, bone mineral density; CI, confidence interval

*p-value < 0.05
