## Supplemental tables for "Pretreatment bone turnover status and long-term bone mineral density response to denosumab"

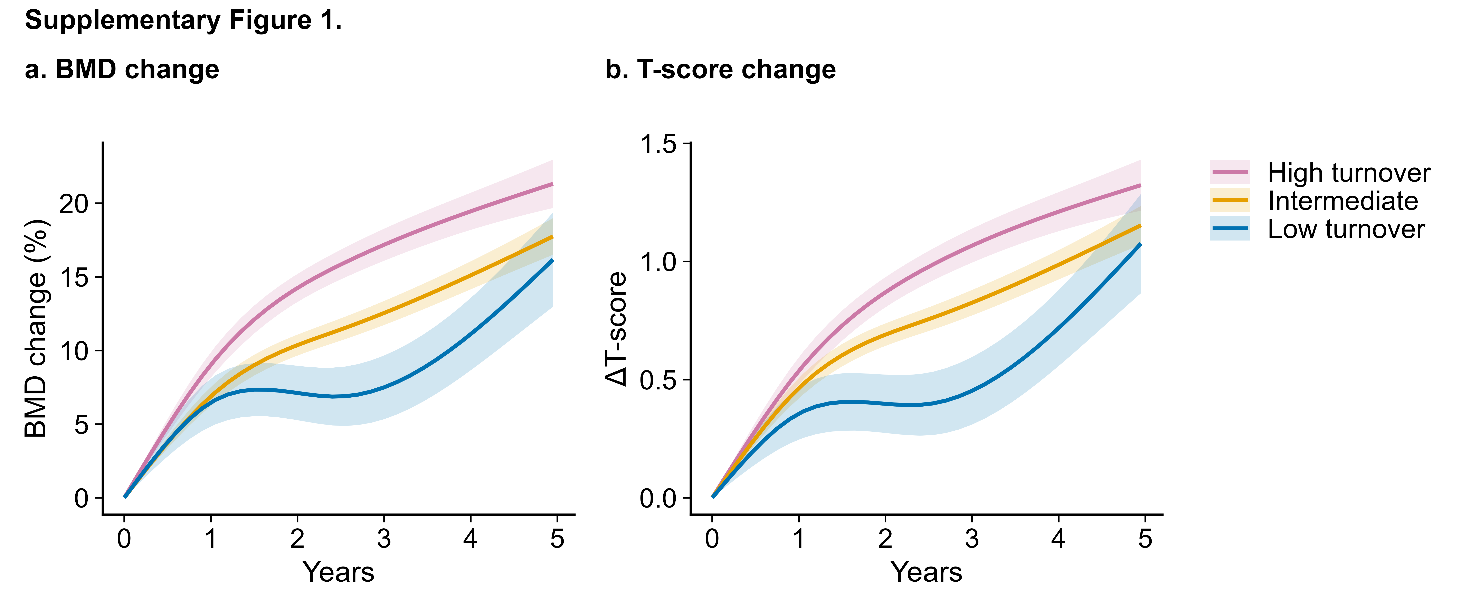

**Supplementary Fig. S1. Adjusted trajectories of lumbar spine DXA outcomes in the treatment-naïve cohort.**
Adjusted trajectories of percent BMD gain from baseline (a) and change in T-score (b) were estimated in the treatment-naïve cohort using multivariable linear mixed-effects models with natural cubic splines for follow-up time. Models were adjusted for age, sex, BMI, eGFR, baseline lumbar spine DXA value or T-score, and type of vitamin D supplementation during denosumab treatment. Shaded areas indicate standard errors.

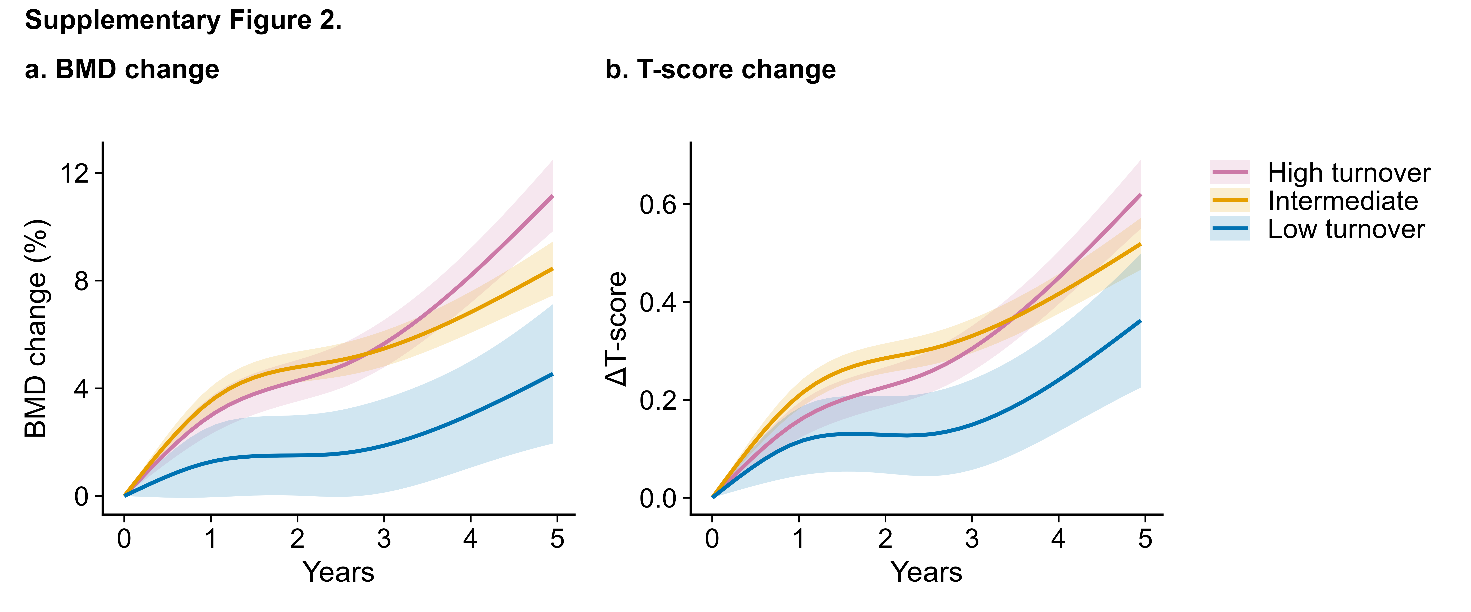

**Supplementary Fig. S2. Adjusted trajectories of total hip DXA outcomes in the treatment-naïve cohort.**
Adjusted trajectories of percent BMD gain from baseline (a) and change in T-score (b) were estimated in the treatment-naïve cohort using multivariable linear mixed-effects models with natural cubic splines for follow-up time. Models were adjusted for age, sex, BMI, eGFR, baseline total hip DXA value or T-score, and type of vitamin D supplementation during denosumab treatment. Shaded areas indicate standard errors.

**Supplementary Table S1. Patient demographic in treatment-naïve patients**

|  | High turnover | Intermediate | Low turnover | p-value |
| --- | --- | --- | --- | --- |
| n | 37 | 64 | 12 |  |
| Age (mean (SD)) | 77.3 (8.4) | 74.6 (8.6) | 79.4 (8.0) | 0.117 |
| Male, n(%) | 4 (10.8) | 6 (9.4) | 0 (0.0) | 0.506 |
| BMI (mean (SD)) | 20.4 (4.8) | 22.3 (3.9) | 24.3 (6.4) | 0.083 |
| Fracture history, n(%) | 30 (81.1) | 38 (59.4) | 6 (50.0) | 0.043* |
| Smoking, n(%) | 2 (5.4) | 1 (1.6) | 0 (0.0) | 0.426 |
| Glucocorticoid use , n(%) | 0 (0.0) | 3 (4.7) | 1 (8.3) | 0.299 |
| eGFR, mL/min/1.73 m², (mean (SD)) | 66.3 (21.6) | 64.1 (16.4) | 53.5 (21.7) | 0.224 |
| Albumin, g/dL, (mean (SD)) | 4.1 (0.3) | 4.2 (0.3) | 4.4 (0.2) | 0.014* |
| Calcium, mg/dL, (mean (SD)) | 9.3 (0.4) | 9.3 (0.3) | 9.5 (0.7) | 0.387 |
| IP, mg/dL, (mean (SD)) | 3.7 (0.5) | 3.6 (0.4) | 3.7 (0.5) | 0.331 |
| ALP, U/L, (mean (SD)) | 355.3 (109.4) | 277.4 (89.3) | 222.2 (66.0) | <0.001* |
| TRACP-5b, mU/dL, (median [IQR]) | 750.0 [629.0, 824.0] | 462.5 [383.2, 568.0] | 231.0 [198.2, 268.0] | <0.001* |
| Total P1NP, µg/L, (median [IQR]) | 106.3 [92.4, 146.6] | 55.5 [47.4, 69.6] | 28.4 [21.6, 29.6] | <0.001* |
| Lumbar spine BMD, g/cm², (mean (SD)) | 0.722 (0.169) | 0.766 (0.157) | 0.793 (0.148) | 0.311 |
| Lumbar spine T-score (mean (SD)) | -2.376 (1.513) | -1.989 (1.404) | -1.717 (1.305) | 0.294 |
| Total hip BMD, g/cm², (mean (SD)) | 0.576 (0.125) | 0.643 (0.120) | 0.678 (0.081) | 0.006* |
| Total hip T-score (mean (SD)) | -2.984 (1.243) | -2.320 (1.209) | -1.983 (0.802) | 0.007* |

BMI, body mass index; VitD, vitamin D supplementation; PTH, parathyroid hormone analog; SERM, selective estrogen receptor modulator; eGFR, estimated glomerular filtration rate; IP, inorganic phosphorus; ALP, alkaline phosphatase; TRACP-5b, tartrate-resistant acid phosphatase 5b; P1NP, N-terminal propeptide of type I procollagen; BMD, bone mineral density; SD, standard deviation; IQR, interquartile range.

*p < 0.05

**Supplementary Table S2. TRACP-5b–specific P1NP thresholds for TI phenotype classification**

| TRACP-5b | P1NP |  |
| --- | --- | --- |
|  | Total P1NP threshold for low-turnover group | Total P1NP threshold for high-turnover group |
| 120 | 105.4 | 575.5 |
| 150 | 80 | 436.9 |
| 200 | 56.1 | 306.2 |
| 300 | 34 | 185.6 |
| 400 | 23.8 | 130.1 |
| 500 | 18.1 | 98.8 |
| 600 | 14.4 | 78.9 |
| 800 | 10.1 | 55.3 |

TRACP-5b, tartrate-resistant acid phosphatase 5b; P1NP, N-terminal propeptide of type I procollagen; TI, turnover index.

**Supplementary Table S3. ANOVA Type III from multivariable mixed-effects models for lumbar spine DXA outcomes**

| Outcome | term | NumDF | DenDF | F value | p-value |
| --- | --- | --- | --- | --- | --- |
| %BMD change | Year | 3 | 478.0 | 273.5 | <0.001* |
|  | Group | 2 | 396.3 | 2.0 | 0.134 |
|  | Year × Group | 6 | 535.0 | 10.2 | <0.001* |
| ΔT-score | Year | 3 | 532.4 | 200.1 | <0.001* |
|  | Group | 2 | 532.8 | 1.2 | 0.299 |
|  | Year × Group | 6 | 586.6 | 6.1 | <0.001* |

Type III analysis of variance was performed using Satterthwaite’s approximation for degrees of freedom. Models included follow-up year, TI phenotype group, and their interaction, and were adjusted for age, sex, BMI, eGFR, baseline lumbar spine DXA value, prior osteoporosis treatment status, and type of vitamin D supplementation during denosumab treatment.

BMD, bone mineral density; DXA, dual-energy X-ray absorptiometry; TI, turnover index; NumDF, numerator degrees of freedom; DenDF, denominator degrees of freedom; BMI, body mass index; eGFR, estimated glomerular filtration rate.

**Supplementary Table S4. Adjusted lumbar spine DXA up to 5-years in treatment-naïve patients**

| Spine BMD |  | Adjusted mean | Difference [95% CI] | Adjusted mean | Difference [95% CI] | Adjusted mean | p-value | | |
| --- | --- | --- | --- | --- | --- | --- | --- | --- | --- |
|  | year | High turnover | H vs I | Intermediate | I vs L | Low turnover | H vs I | I vs L | H vs L |
| %BMD change | 1 | 9.03 (0.86) | 2.14 [0.05, 4.24] | 6.89 (0.64) | 0.41 [-3.01, 3.84] | 6.47 (1.62) | 0.134 | 0.812 | 0.329 |
|  | 2 | 14.25 (0.95) | 3.88 [1.56, 6.20] | 10.37 (0.71) | 3.25 [-0.63, 7.14] | 7.11 (1.84) | 0.002* | 0.100 | 0.002* |
|  | 5 | 21.42 (1.67) | 3.53 [-0.57, 7.63] | 17.89 (1.26) | 1.43 [-5.47, 8.34] | 16.45 (3.25) | 0.273 | 0.682 | 0.355 |
| ΔT-score | 1 | 0.537 (0.058) | 0.073 [-0.068, 0.214] | 0.464 (0.043) | 0.108 [-0.124, 0.339] | 0.356 (0.110) | 0.623 | 0.623 | 0.437 |
|  | 2 | 0.870 (0.064) | 0.180 [0.024, 0.335] | 0.691 (0.048) | 0.293 [0.033, 0.553] | 0.398 (0.123) | 0.047* | 0.047* | 0.002* |
|  | 5 | 1.329 (0.110) | 0.166 [-0.104, 0.437] | 1.163 (0.083) | 0.067 [-0.387, 0.520] | 1.096 (0.214) | 0.676 | 0.772 | 0.676 |

Values are adjusted estimated marginal means with standard errors derived from multivariable linear mixed-effects models. Models were adjusted for age, sex, BMI, eGFR, baseline DXA value at the lumbar spine, and type of vitamin D supplementation during denosumab treatment. Differences are presented with unadjusted 95% confidence intervals. P values were adjusted using the Holm method for the three pairwise comparisons within each time point.

BMD, bone mineral density; CI, confidence interval

*p-value < 0.05

**Supplementary Table S5. ANOVA Type III from multivariable mixed-effects models for total hip DXA outcomes**

| Outcome | term | NumDF | DenDF | F value | p-value |
| --- | --- | --- | --- | --- | --- |
| %BMD change | Year | 3 | 482.2 | 45.73 | <0.001* |
|  | Group | 2 | 612.2 | 0.38 | 0.677 |
|  | Year × Group | 6 | 555.0 | 5.33 | <0.001* |
| ΔT-score | Year | 3 | 502.2 | 55.22 | <0.001* |
|  | Group | 2 | 548.0 | 0.26 | 0.774 |
|  | Year × Group | 6 | 503.7 | 4.13 | 0.001* |

Type III analysis of variance was performed using Satterthwaite’s approximation for degrees of freedom. Models included follow-up year, TI phenotype group, and their interaction, and were adjusted for age, sex, BMI, eGFR, baseline total hip DXA value, prior osteoporosis treatment status, and type of vitamin D supplementation during denosumab treatment.

BMD, bone mineral density; DXA, dual-energy X-ray absorptiometry; TI, turnover index; NumDF, numerator degrees of freedom; DenDF, denominator degrees of freedom; BMI, body mass index; eGFR, estimated glomerular filtration rate.

**Supplementary Table S6. Adjusted total hip DXA up to 5-years in treatment-naïve patients**

| Total hip BMD |  | Adjusted mean | Difference [95% CI] | Adjusted mean | Difference [95% CI] | Adjusted mean | p-value | | |
| --- | --- | --- | --- | --- | --- | --- | --- | --- | --- |
|  | year | High turnover | H vs I | Intermediate | I vs L | Low turnover | H vs I | I vs L | H vs L |
| %BMD change | 1 | 2.99 (0.69) | -0.55 [-2.24, 1.15] | 3.54 (0.52) | 2.27 [-0.52, 5.05] | 1.27 (1.32) | 0.529 | 0.333 | 0.499 |
|  | 2 | 4.28 (0.77) | -0.50 [-2.38, 1.37] | 4.79 (0.58) | 3.28 [0.14, 6.42] | 1.51 (1.49) | 0.598 | 0.123 | 0.198 |
|  | 5 | 11.33 (1.35) | 2.79 [-0.52, 6.10] | 8.54 (1.02) | 3.92 [-1.66, 9.50] | 4.62 (2.63) | 0.197 | 0.197 | 0.075 |
| ΔT-score | 1 | 0.158 (0.036) | -0.051 [-0.140, 0.038] | 0.209 (0.027) | 0.094 [-0.052, 0.240] | 0.114 (0.069) | 0.615 | 0.615 | 0.615 |
|  | 2 | 0.227 (0.040) | -0.059 [-0.157, 0.040] | 0.285 (0.030) | 0.157 [-0.009, 0.323] | 0.128 (0.079) | 0.489 | 0.191 | 0.489 |
|  | 5 | 0.630 (0.071) | 0.105 [-0.070, 0.280] | 0.525 (0.054) | 0.155 [-0.140, 0.451] | 0.369 (0.139) | 0.474 | 0.474 | 0.295 |

Values are adjusted estimated marginal means with standard errors derived from multivariable linear mixed-effects models. Models were adjusted for age, sex, BMI, eGFR, baseline DXA value at the total hip, and type of vitamin D supplementation during denosumab treatment. Differences are presented with unadjusted 95% confidence intervals. P values were adjusted using the Holm method for the three pairwise comparisons within each time point.

BMD, bone mineral density; CI, confidence interval

*p-value < 0.05

**Supplementary Table S7. Mortality and fracture events during denosumab treatment**

|  | High turnover | Intermediate | Low turnover | p-value |
| --- | --- | --- | --- | --- |
| n | 52 | 114 | 64 |  |
| Death, n, n(%) | 8 (15.4) | 15 (13.2) | 6 (9.4) | 0.61 |
| Reasons, n(%) |  |  |  |  |
| Cerebral vascular event | 0 (0.0) | 2 (1.8) | 0 (0.0) |  |
| Malignancy | 2 (3.8) | 1 (0.9) | 1 (1.5) |  |
| Gastrointestinal event | 0 (0.0) | 2 (1.8) | 0 (0.0) |  |
| Septic shock | 1 (1.9) | 0 (0.0) | 0 (0.0) |  |
| Cardiovascular event | 2 (3.8) | 5 (4.4) | 0 (0.0) |  |
| Multiple organ failure | 0 (0.0) | 1 (0.9) | 0 (0.0) |  |
| Pneumonia | 3 (5.7) | 2 (1.8) | 2 (3.1) |  |
| Respiratory failure | 0 (0.0) | 1 (0.9) | 0 (0.0) |  |
| Empyema | 0 (0.0) | 0 (0.0) | 1 (1.6) |  |
| Renal failure | 0 (0.0) | 0 (0.0) | 1 (1.6) |  |
| Unknown reason | 0 (0.0) | 1 (0.9) | 1 (1.6) |  |
| Number of fracture during follow-up, n(%) | |  |  | 0.99 |
| 0 | 44 (84.6) | 93 (81.6) | 53 (82.8) |  |
| 1 | 7 (13.5) | 19 (16.7) | 10 (15.6) |  |
| 2 | 1 (1.9) | 2 (1.8) | 1 (1.6) |  |
| Major osteoporotic fracture, n, n(%) | 4 (7.7) | 10 (8.8) | 7 (10.9) | 0.82 |
